# Understanding Post-Acute Sequelae of SARS-CoV-2 Infection through Data-Driven Analysis with Longitudinal Electronic Health Records: Findings from the RECOVER Initiative

**DOI:** 10.1101/2022.05.21.22275420

**Authors:** Chengxi Zang, Yongkang Zhang, Jie Xu, Jiang Bian, Dmitry Morozyuk, Edward J. Schenck, Dhruv Khullar, Anna S. Nordvig, Elizabeth A. Shenkman, Russel L. Rothman, Jason P. Block, Kristin Lyman, Mark Weiner, Thomas W. Carton, Fei Wang, Rainu Kaushal

**Affiliations:** Department of Population Health Sciences, Weill Cornell Medicine, New York, NY, USA; Department of Health Outcomes Biomedical Informatics, University of Florida, Gainesville, FL, USA; Division of Pulmonary and Critical Care Medicine, Weill Cornell Department of Medicine, New York, NY, USA; Department of Neurology, Weill Cornell Medicine, New York, NY, USA; Center for Health Services Research, Vanderbilt University Medical Center, Nashville, Tennessee, USA; Department of Population Medicine, Harvard Pilgrim Health Care Institute, Harvard Medical School, Boston, MA, USA; Louisiana Public Health Institute, New Orleans, Louisiana, USA

## Abstract

Recent studies have investigated post-acute sequelae of SARS-CoV-2 infection (PASC) using real-world patient data such as electronic health records (EHR). Prior studies have typically been conducted on patient cohorts with small sample sizes^1^ or specific patient populations^2,3^ limiting generalizability. This study aims to characterize PASC using the EHR data warehouses from two large national patient-centered clinical research networks (PCORnet), INSIGHT and OneFlorida+, which include 11 million patients in New York City (NYC) and 16.8 million patients in Florida respectively. With a high-throughput causal inference pipeline using high-dimensional inverse propensity score adjustment, we identified a broad list of diagnoses and medications with significantly higher incidence 30-180 days after the laboratory-confirmed SARS-CoV-2 infection compared to non-infected patients. We found more PASC diagnoses and a higher risk of PASC in NYC than in Florida, which highlights the heterogeneity of PASC in different populations.

## Introduction

The global COVID-19 pandemic from late 2019 has led to more than 434 million infections and 5.9 million deaths as of March 1, 2022^4^. Growing scientific and clinical evidence has demonstrated potential post-acute and long-term effects of SARS-CoV-2 infection in multiple organ systems^5^, including cardiovascular^6^, mental health^7^, neurological^8^, and metabolic^9^ among other systems. Recently, several retrospective cohort analyses have described post-acute sequelae of SARS-CoV-2 infection (PASC) using real-world patient data^2,10,11^. These studies typically start with a predefined list of PASC symptoms and signs, and then contrast their incidence or burden in SARS-CoV-2 positive patients versus appropriate controls. Different analytical pipelines have been utilized, such as causal inference^2^, regression analysis^12^, and network analysis^13^. There are two major challenges to these existing studies. First, the disease etiology and pathophysiology of PASC is complicated, and our current state of knowledge is still far from complete. This means that conventional hypothesis-driven study design may miss potential PASC symptoms and signs or result in biased findings. Second, prior studies have typically been conducted on patient cohorts with small sample sizes^1^, or specific patient populations^2,3^. It is unclear how generalizable the results are from these studies when applied to the general patient population, and how PASC varies over broad patient populations with different characteristics.

In this study, we aim to address these two challenges by developing a high-throughput causal inference pipeline to identify potential PASC symptoms and signs using electronic health records (EHR) from two large national patient-centered clinical research networks (PCORnet)^14^: the INSIGHT network^15^ covering patients in the New York City (NYC) metropolitan area and the OneFlorida+ network^16^ covering patients from Florida, Georgia and Alabama. We started with a broad list of 137 potential PASC diagnoses and 459 potential PASC medications (See Method for the construction of both the diagnosis and medication lists). For each of diagnoses or medications, we built an outcome-specific cohort with patients who are free of it at baseline, applied inverse propensity treatment re-weighting (IPTW) to adjust for high-dimensional hypothetical confounders collected from the baseline period, and calculated its adjusted hazard ratio and excess burden in the post-acute phase of the SARS-CoV-2 infection compared to non-infected patients (See an illustration in Fig. 1 and details in the Method section). We only focused on new incidences in the post-acute period in this study because it provided a clean way of defining PASC phenotypes without complicated consideration of pre-existing conditions. 25 diagnosis categories and 51 medications involving a wide range of organ systems were identified to be associated with SARS-CoV-2 exposure from the INSIGHT cohort. However, by applying the same methodology for the OneFlorida+ cohort, we only found 9 diagnosis categories and 9 medications, and they were subset of the findings from INSIGHT. This discrepancy highlights the heterogeneity of PASC and the need for replication studies over different populations before robust conclusions about PASC can be made. This study is part of the NIH Researching COVID to Enhance Recovery (RECOVER) Initiative, which seeks to understand, treat, and prevent the post-acute sequelae of SARS-CoV-2 infection (PASC). For more information on RECOVER, visit https://recovercovid.org/.

**Fig. 1.**
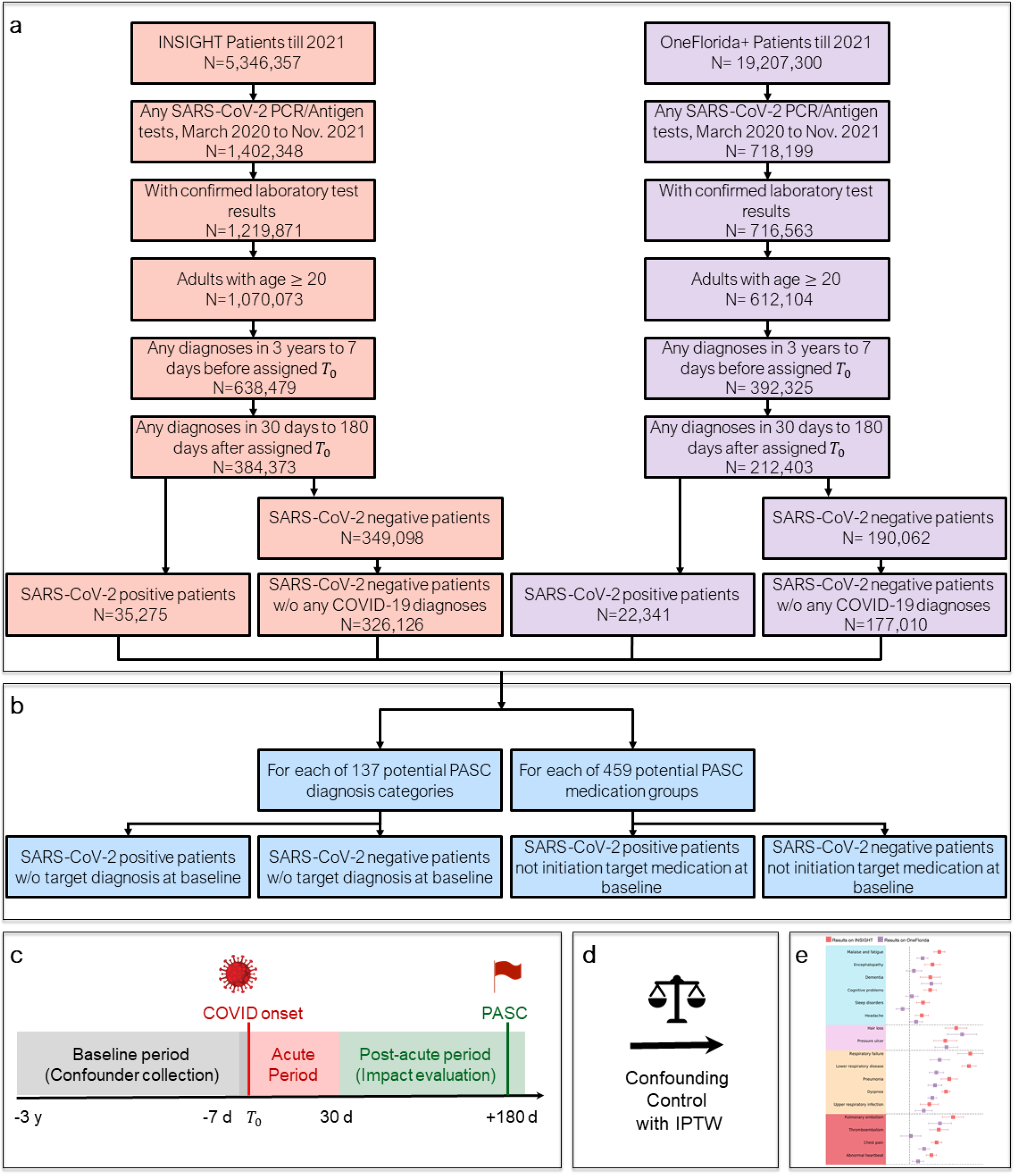
Overall data-driven high-throughput screening framework for the post-acute sequelae of SARS-CoV-2 infection (PASC) in both the INSIGHT and OneFlorida+ cohorts, March 2020 to November 2021. a. Selection of patients from the INSIGHT and OneFlorida+ EHR warehouses. b. High-throughput construction of PASC-specific case and control groups that patients did not have target condition at baseline. c. Study design. The PASC outcomes were ascertained from day 30 after the SARS-CoV-2 infection and the adjusted risk were computed at 180 days after the SARS-CoV-2 infection. d. Confounding control with inverse propensity treatment re-weighting (IPTW). e. Likely PASC were identified in the INSIGHT and OneFlorida+ cohorts respectively. Identified PASC were compared across the two cohorts.

## Results

### Population statistics

We identified potential PASC conditions using two different cohorts. The first cohort was built from the INSIGHT network^15^, which contained 35,275 adult patients (age ≥ 20) with lab-confirmed SARS-CoV-2 infection who survived the first 30 days of infection from March 2020 to November 2021 in NYC and 326,126 eligible non-infected controls. Our second cohort was built from the OneFlorida+ network^16^ with 22,341 eligible lab-confirmed SARS-CoV-2 positive patients who survived the first 30 days of infection during the same period in Florida, Georgia, and Alabama and 177,010 non-infected controls. To ensure that patients were connected to healthcare systems (and thus available for observation before and after their index encounters), we required eligible patients to have at least one diagnosis record within three years to one week prior to the index date and at least one diagnosis record within 30 days to 180 days after the index date. We also required no COVID-19-related diagnoses for the control patients (see Methods for the definitions of index date and lab confirmations, and Fig. 1 for inclusion-exclusion cascade). We identified new-onset diagnoses and medications for SARS-CoV-2 infected patients in excess of control patients 30-180 days after the index date as potential PASC conditions.

We summarized the baseline characteristics of both the INSIGHT cohort and OneFlorida+ cohort in Table 1 from information that was available on patients in clinical data; demographic information was collected from patients when they registered for care within the healthcare systems. We observed significant differences between the two cohorts regarding age, gender, race, area deprivation index, and outbreak waves. The INSIGHT cohort contained SARS-CoV-2 infected patients mainly from the New York metropolitan area with the median area deprivation index (ADI)^17^ 15 (6-24) in the SARS-CoV-2 infected patient group, indicating fewer disadvantaged neighborhoods than the OneFlorida+ cohort whose median ADI was 58 (41-76). Indeed, the OneFlorida+ cohort consisted of a mixture of urban, sub-urban and rural populations in Florida and selected cities in Georgia and Alabama (see Methods). The median age of SARS-CoV-2 infected patients in the INSIGHT cohort was 55 (38-68), older than the OneFlorida+ cohort with median age of 50 (34-64). Plus, more female SARS-CoV-2 infected patients were in the OneFlorida+ cohort (62.7%) than in the INSIGHT cohort (58.6%). The INSIGHT cohort also had a more diverse population with 34.7% white and 54.9% others (Asian and others including American Indian or Alaska Native, Native Hawaiian or other Pacific Islander, multiple races, etc.); the OneFlorida+ cohort had a majority of patients identifying as White race (51.0%). Additionally, there is a higher proportion of patients infected early in the pandemic in the INSIGHT cohort (31.8% of all infected patients were from March 2020 to June 2020) compared to the OneFlorida+ cohort (9.1% of cases were from March 2020 to June 2020). Different temporal patterns of new cases per month across two cohorts are illustrated in Extended Data Fig. 1. The two networks also differed in care settings connected to patient encounters and treatments utilized for infected patients (e.g., more inpatient visits and more prescriptions of corticosteroids in the OneFlorida+ cohort than in the INSIGHT cohort).

**Table 1.** Baseline characteristics of the lab-confirmed SARS-CoV-2 Positive patients and SARS-CoV-2 Negative patients in the INSIGHT and OneFlorida+ cohorts, March 2020 to November 2021^a^.

|  | INSIGHT |  |  | OneFlorida+ |  |  |
| --- | --- | --- | --- | --- | --- | --- |
| Characteristics | SARS-CoV-2 Positive (N=35,275) | SARS-CoV-2 Negative (N=326,126) | SMD<sup>b</sup> | SARS-CoV-2 Positive (N=22,341) | SARS-CoV-2 Negative (N=177,010) | SMD |
| Median age (IQR) — yrs | 55 (38-68) | 57 (40-69) | -0.09 | 50 (34-64) | 57 (40-69) | -0.27 |
| **Age group — no. (%)** |  |  |  |  |  |  |
| 20-<40 years | 9,529 (27.0) | 77,403 (23.7) | 0.08 | 7,506 (33.6) | 42,286 (23.9) | 0.22 |
| 40-<55 years | 7,975 (22.6) | 70,313 (21.6) | 0.03 | 5,473 (24.5) | 37,555 (21.2) | 0.08 |
| 55-<65 years | 6,965 (19.7) | 66,361 (20.3) | -0.02 | 4,036 (18.1) | 37,142 (21.0) | -0.07 |
| 65-<75 years | 5,712 (16.2) | 62,860 (19.3) | -0.08 | 2,929 (13.1) | 34,601 (19.5) | -0.17 |
| 75+ years | 5,094 (14.4) | 49,189 (15.1) | -0.02 | 2,397 (10.7) | 25,426 (14.4) | -0.11 |
| **Sex — no. (%)** |  |  |  |  |  |  |
| Female | 20,686 (58.6) | 196,730 (60.3) | -0.03 | 14,004 (62.7) | 106,963 (60.4) | 0.05 |
| Male | 14,586 (41.3) | 129,360 (39.7) | 0.03 | 8,335 (37.3) | 70,034 (39.6) | -0.05 |
| **Race — no. (%)** |  |  |  |  |  |  |
| Asian | 1,736 (4.9) | 17,439 (5.3) | -0.02 | 275 (1.2) | 2,912 (1.6) | -0.03 |
| Black | 7,791 (22.1) | 62,281 (19.1) | 0.07 | 6,504 (29.1) | 35,381 (20.0) | 0.21 |
| White | 12,233 (34.7) | 139,512 (42.8) | -0.17 | 11,398 (51.0) | 105,521 (59.6) | -0.17 |
| Other | 9,844 (27.9) | 69,406 (21.3) | 0.15 | 3,730 (16.7) | 30,138 (17.0) | -0.01 |
| Unknown | 3,671 (10.4) | 37,488 (11.5) | -0.03 | 434 (1.9) | 3,058 (1.7) | 0.02 |
| **Ethnic group — no. (%)** |  |  |  |  |  |  |
| Hispanic | 10,658 (30.2) | 73,522 (22.5) | 0.17 | 4,500 (20.1) | 21,484 (12.1) | 0.22 |
| Not Hispanic | 20,838 (59.1) | 216,179 (66.3) | -0.15 | 14,798 (66.2) | 120,315 (68.0) | -0.04 |
| Unknown | 3,779 (10.7) | 36,425 (11.2) | -0.01 | 3,043 (13.6) | 35,211 (19.9) | -0.17 |
| Median ADI (IQR) — rank | 15 (6-24) | 13 (5-23) | 0.03 | 58 (41-76) | 53 (36-72) | 0.19 |
| **Hospital visits in the past 3 years — no. (%)** |  |  |  |  |  |  |
| Inpatient 0 | 25,717 (72.9) | 278,784 (85.5) | -0.31 | 12,838 (57.5) | 112,480 (63.5) | -0.12 |
| Inpatient 1-2 | 6,805 (19.3) | 37,297 (11.4) | 0.22 | 4,614 (20.7) | 33,658 (19.0) | 0.04 |
| Inpatient ≥3 | 2,753 (7.8) | 10,045 (3.1) | 0.21 | 4,889 (21.9) | 30,872 (17.4) | 0.11 |
| Outpatient 0 | 1,570 (4.5) | 8,956 (2.7) | 0.09 | 3,757 (16.8) | 19,163 (10.8) | 0.17 |
| Outpatient 1-2 | 3,327 (9.4) | 32,047 (9.8) | -0.01 | 2,617 (11.7) | 18,622 (10.5) | 0.04 |
| Outpatient ≥3 | 30,378 (86.1) | 285,123 (87.4) | -0.04 | 15,967 (71.5) | 139,225 (78.7) | -0.17 |
| Emergency 0 | 20,351 (57.7) | 238,527 (73.1) | -0.33 | 8,883 (39.8) | 96,496 (54.5) | -0.30 |
| Emergency 1-2 | 9,518 (27.0) | 61,973 (19.0) | 0.19 | 5,151 (23.1) | 35,094 (19.8) | 0.08 |
| Emergency >=3 | 5,406 (15.3) | 25,626 (7.9) | 0.23 | 8,307 (37.2) | 45,420 (25.7) | 0.25 |
| **BMI kg/m<sup>2</sup> (IQR)** | 27 (21-32) | 25 (1-30) | 0.02 | 30 (25-35) | 28 (24-34) | 0.00 |
| BMI >=30 obese | 9,751 (27.6) | 74,602 (22.9) | 0.11 | 8,497 (38.0) | 62,764 (35.5) | 0.05 |
| **Index time period — no. (%)** |  |  |  |  |  |  |
| 03/20-06/20 | 11,235 (31.8) | 53,988 (16.6) | 0.36 | 2,032 (9.1) | 37,363 (21.1) | -0.34 |
| 07/20-10/20 | 2,018 (5.7) | 111,409 (34.2) | -0.76 | 6,035 (27.0) | 54,060 (30.5) | -0.08 |
| 11/20-02/21 | 14,637 (41.5) | 88,009 (27.0) | 0.31 | 6,254 (28.0) | 38,536 (21.8) | 0.14 |
| 03/21-06/21 | 5,573 (15.8) | 54,234 (16.6) | -0.02 | 2,315 (10.4) | 27,985 (15.8) | -0.16 |
| 07/21-11/21 | 1,812 (5.1) | 18,486 (5.7) | -0.02 | 5,705 (25.5) | 19,066 (10.8) | 0.39 |
| **Pre-existing conditions — no. (%)<sup>c</sup>** |  |  |  |  |  |  |
| Alcohol Abuse | 1,018 (2.9) | 7,689 (2.4) | 0.03 | 723 (3.2) | 7,547 (4.3) | -0.05 |
| Anemia | 4,862 (13.8) | 27,588 (8.5) | 0.17 | 4,071 (18.2) | 26,085 (14.7) | 0.09 |
| Arrhythmia | 5,350 (15.2) | 33,729 (10.3) | 0.14 | 3,075 (13.8) | 24,187 (13.7) | 0.00 |
| Asthma | 3,950 (11.2) | 23,852 (7.3) | 0.13 | 2,596 (11.6) | 15,679 (8.9) | 0.09 |
| Autism | 47 (0.1) | 288 (0.1) | 0.01 | 89 (0.4) | 578 (0.3) | 0.01 |
| Cancer | 3,616 (10.3) | 34,169 (10.5) | -0.01 | 1,968 (8.8) | 24,141 (13.6) | -0.15 |
| Chronic Kidney Disease | 5,126 (14.5) | 24,720 (7.6) | 0.22 | 3,257 (14.6) | 22,656 (12.8) | 0.05 |
| Chronic Pulmonary Disorders | 6,209 (17.6) | 38,302 (11.7) | 0.17 | 4,435 (19.9) | 31,803 (18.0) | 0.05 |
| Cirrhosis | 582 (1.6) | 4,097 (1.3) | 0.03 | 368 (1.6) | 3,856 (2.2) | -0.04 |
| Coagulopathy | 2,511 (7.1) | 9,621 (3.0) | 0.19 | 1,141 (5.1) | 7,418 (4.2) | 0.04 |
| Congestive Heart Failure | 3,682 (10.4) | 21,376 (6.6) | 0.14 | 2,920 (13.1) | 20,631 (11.7) | 0.04 |
| COPD | 1,726 (4.9) | 9,644 (3.0) | 0.10 | 1,600 (7.2) | 13,474 (7.6) | -0.02 |
| Coronary Artery Disease | 4,658 (13.2) | 30,523 (9.4) | 0.12 | 2,742 (12.3) | 22,397 (12.7) | -0.01 |
| Cystic Fibrosis | 13 (0.0) | 148 (0.0) | 0.00 | 16 (0.1) | 159 (0.1) | -0.01 |
| Dementia | 1,404 (4.0) | 5,330 (1.6) | 0.14 | 909 (4.1) | 4,100 (2.3) | 0.10 |
| Diabetes Type 1 | 435 (1.2) | 2,262 (0.7) | 0.06 | 394 (1.8) | 2,414 (1.4) | 0.03 |
| Diabetes Type 2 | 7,681 (21.8) | 43,233 (13.3) | 0.23 | 5,323 (23.8) | 34,512 (19.5) | 0.11 |
| Down's Syndrome | 30 (0.1) | 105 (0.0) | 0.02 | 35 (0.2) | 146 (0.1) | 0.02 |
| End Stage Renal Disease on Dialysis | 1,573 (4.5) | 5,701 (1.7) | 0.16 | 895 (4.0) | 5,094 (2.9) | 0.06 |
| Hemiplegia | 440 (1.2) | 2,242 (0.7) | 0.06 | 345 (1.5) | 2,050 (1.2) | 0.03 |
| HIV | 586 (1.7) | 5,473 (1.7) | 0.00 | 188 (0.8) | 1,601 (0.9) | -0.01 |
| Hypertension | 13,796 (39.1) | 95,865 (29.4) | 0.21 | 9,854 (44.1) | 73,739 (41.7) | 0.05 |
| Inflammatory Bowel Disorder | 367 (1.0) | 4,994 (1.5) | -0.04 | 251 (1.1) | 2,699 (1.5) | -0.04 |
| Lupus or Systemic Lupus Erythematosus | 290 (0.8) | 1,894 (0.6) | 0.03 | 257 (1.2) | 1,536 (0.9) | 0.03 |
| Mental Health Disorders | 3,682 (10.4) | 24,530 (7.5) | 0.10 | 3,983 (17.8) | 29,640 (16.7) | 0.03 |
| Multiple Sclerosis | 187 (0.5) | 1,224 (0.4) | 0.02 | 109 (0.5) | 779 (0.4) | 0.01 |
| Other Substance Abuse | 2,225 (6.3) | 18,661 (5.7) | 0.02 | 2,694 (12.1) | 26,706 (15.1) | -0.09 |
| Parkinson's Disease | 222 (0.6) | 1,781 (0.5) | 0.01 | 143 (0.6) | 1,566 (0.9) | -0.03 |
| Peripheral vascular disorders | 2,562 (7.3) | 16,914 (5.2) | 0.09 | 1,695 (7.6) | 13,416 (7.6) | 0.00 |
| Pregnant | 1,608 (4.6) | 15,722 (4.8) | -0.01 | 1,504 (6.7) | 10,350 (5.8) | 0.04 |
| Pulmonary Circulation Disorder | 1,345 (3.8) | 6,206 (1.9) | 0.11 | 927 (4.1) | 5,761 (3.3) | 0.05 |
| Rheumatoid Arthritis | 544 (1.5) | 3,649 (1.1) | 0.04 | 453 (2.0) | 3,221 (1.8) | 0.02 |
| Seizure/Epilepsy | 790 (2.2) | 4,596 (1.4) | 0.06 | 795 (3.6) | 5,124 (2.9) | 0.04 |
| Severe Obesity (BMI≥40 kg/m<sup>2</sup>) | 2,519 (7.1) | 16,613 (5.1) | 0.09 | 2,779 (12.4) | 15,006 (8.5) | 0.13 |
| Sickle Cell | 255 (0.7) | 1,536 (0.5) | 0.03 | 227 (1.0) | 1,108 (0.6) | 0.04 |
| Weight Loss | 1,603 (4.5) | 7,190 (2.2) | 0.13 | 1,125 (5.0) | 7,742 (4.4) | 0.03 |
| Corticosteroids Prescriptions | 4,999 (14.2) | 28,915 (8.9) | 0.17 | 4,253 (19.0) | 27,783 (15.7) | 0.09 |
| Immunosuppressant Prescriptions | 2,110 (6.0) | 10,761 (3.3) | 0.13 | 1,013 (4.5) | 7,281 (4.1) | 0.02 |

a. The lab-confirmed SARS-CoV-2 positive and negative patients were identified by polymerase chain reaction (PCR) test or antigen test. Negative patients were further required no documented COVID-19 related diagnoses at any time. IQR denotes inter-quartile range. Percentage may not sum up to 100 because of rounding.
b. A standardized mean difference (SMD) of $>0.10$ or $<-0.10$ indicates an important effect size difference between the two samples, otherwise, no significant difference is assumed.
c. Coexisting conditions existed if two records in the 3-years prior to index event. See detailed phenotyping codes in the appendix. SLE: Systemic Lupus Erythematosus; COPD: Chronic obstructive pulmonary disease.

### Identified potential PASC conditions in the INSIGHT cohort

We started with a list of 137 potentially PASC-related diagnostic groups defined by ICD-10 diagnosis codes and CCSR categories (Supplementary Table 2) and 359 classes of medications grouped by their active ingredients (See Method) to screen for potential PASC conditions. For each of these diagnoses or medications, we built a specific cohort who didn’t have the condition at baseline (Fig. 1) and conducted a causal inference procedure following the pipeline in Extended Data Table 2 (details provided in Method section) to estimate its risk in the post-acute period of SARS-CoV-2 infected patients compared to non-infected controls. Figure 2 summarizes potential PASC diagnoses (Fig. 2a) and medications (Fig. 2b) identified from the INSIGHT cohort, spanning a broad range of organ systems. We reported their incident risks in the adjusted hazard ratio (Fig. 2) and excess burdens in the adjusted excess cumulative incidence (Fig.3).

**Fig. 2.**
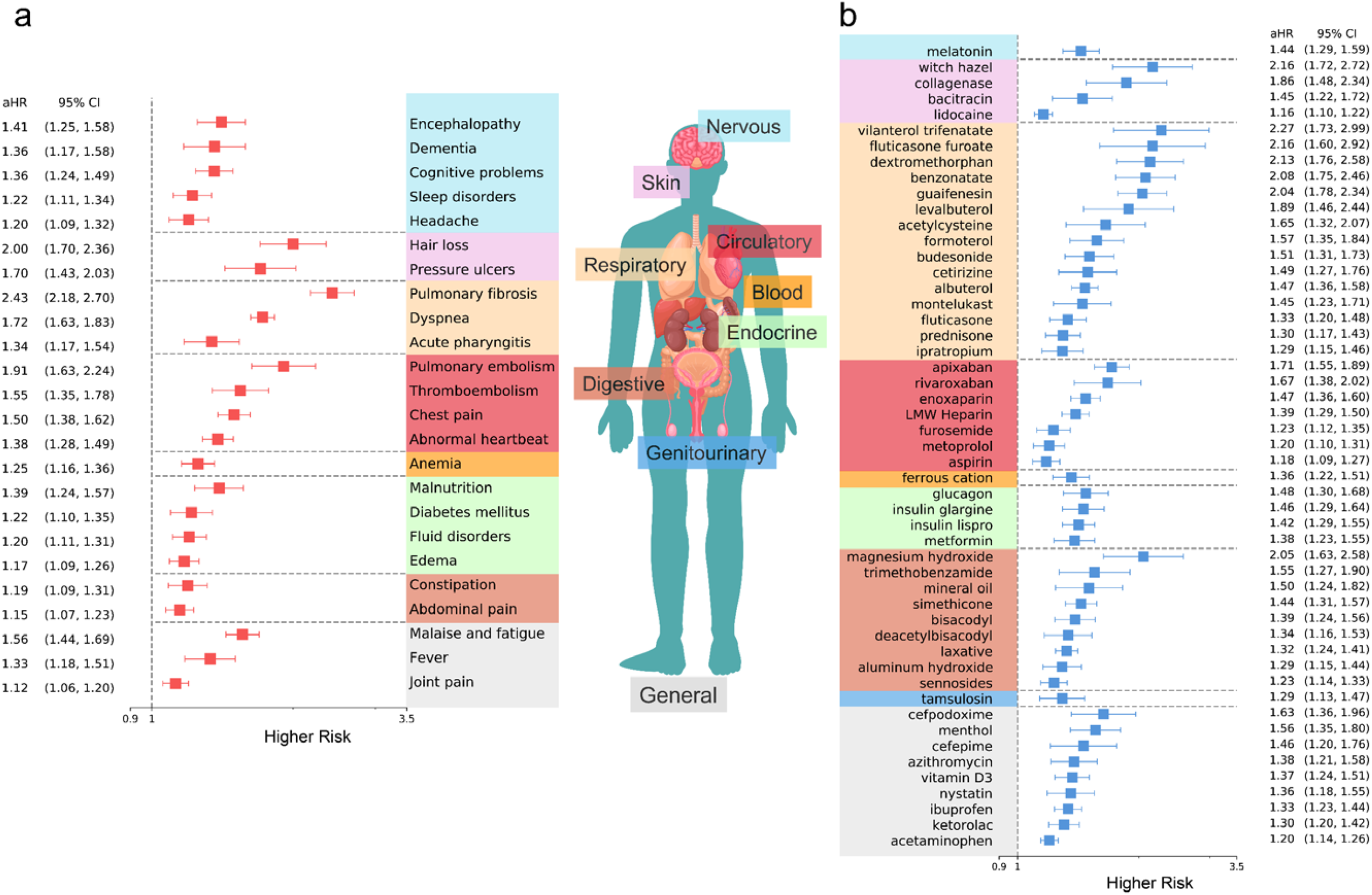
Identified potential incident PASC conditions from the INSIGHT cohort, March 2020 to November 2021. a. The adjusted hazard ratios of incident diagnoses. b. The adjusted hazard ratios of incident use of medications. The sequelae outcomes were ascertained from day 30 after the SARS-CoV-2 infection and the adjusted hazard ratio were computed at 180 days after the SARS-CoV-2 infection. The colors represent different organ systems.

### Nervous System

We observed a number of neurologic conditions which exhibited higher risk in SARS-CoV-2 infected patients after acute infection, including encephalopathy (1.41 [95% CI, 1.25-1.58]), dementia (1.36 [95% CI, 1.17-1.58]), cognitive problems (1.36 [95% CI, 1.24-1.49]), sleep disorders (1.22 [95% CI, 1.11-1.34]), and headache (1.20 [95% CI, 1.09-1.32]). Besides, the insomnia drug melatonin also showed a significantly higher risk of use (1.44 [95% CI, 1.29-1.59]), in line with diagnoses of sleep disorders.

### Skin

Certain skin symptoms also showed significantly higher risk in the post-acute period, including hair loss (2.00 [95% CI, 1.70-2.36]) and pressure ulcers (1.71 [95% CI, 1.44-2.04]), coupled with relevant medications including witch hazel, collagenase, and bacitracin.

### Respiratory System

Several pulmonary manifestations in the post-acute phase were significant. These included pulmonary fibrosis (2.43 [95% CI, 2.18, 2.70]), dyspnea (1.72 [95% CI, 1.63, 1.83]), and acute pharyngitis (1.34 [95% CI, 1.17-1.54]). Besides, a large number of medications in line with these diagnoses also showed significantly higher use, such as asthma or chronic obstructive pulmonary disease drugs (e.g., vilanterol, fluticasone, budesonide, levalbuterol, formoterol, etc.) and cough suppressants (e.g., dextromethorphan, benzonatate, guaifenesin, etc.).

### Circulatory and Blood

Identified cardiovascular manifestations with a higher risk in the post-acute period were pulmonary embolism (1.92 [95% CI, 1.64-2.25]), thromboembolism (1.55 [95% CI, 1.35-1.78]), chest pain (1.50 [95% CI, 1.38-1.62]), and abnormal heartbeat (1.38 [95% CI, 1.28-1.49]), coupled with anticoagulant medications (e.g., apixaban, rivaroxaban, enoxaparin, heparin, etc.) and a beta-blocker metoprolol. We also observed higher risk of anemia (1.25 (95% CI, 1.16-1.36) and ferric cation use (1.36 (95% CI, 1.22-1.51)) in the post-acute phase.

### Endocrine

dentified endocrine, nutritional and metabolic disorders with higher risk were malnutrition (1.39 [95% CI, 1.24-1.57]), diabetes mellitus (1.22 [95% CI, 1.10-1.35]), fluid and electrolyte disorders (1.20 [95% CI, 1.11-1.31]), and edema (1.17 [95% CI, 1.09-1.26]), coupled with higher use of glucagon, insulin, and metformin.

### Digestive System

Digestive system conditions with higher risks were constipation (1.19 [95% CI, 1.09-1.31]) and abdominal pain (1.15 [95% CI, 1.07-1.23]). The associated medications included magnesium hydroxide, trimethobenzamide, and simethicone.

### General and Musculoskeletal

General symptoms including malaise and fatigue (1.56 [95% CI, 1.44-1.69]), fever (1.33 [95% CI 1.18-1.51]), and joint pain (1.12 [95% CI, 1.06-1.20]) showed significantly higher risk, coupled with higher risk of using menthol, ibuprofen, ketorolac, and acetaminophen.

### Stratified Analysis

To better understand the heterogeneity of potential PASC conditions, we conducted a comprehensive stratified analysis to examine how identified diagnoses vary across demographic (age, gender, race) groups, baseline pre-existing conditions, and disease severity in the acute phase operationalized by healthcare utilization (outpatient versus inpatient). We also studied the population that had no documented pre-existing conditions or PASC-like symptoms at baseline, referred to as the healthy population. We estimated the adjusted cumulative incidence of each potential PASC diagnosis per 1,000 patients at 180 days in different groups and compared the excess burden of it^18^, which is the difference between the adjusted cumulative incidences of a specific diagnosis in the SARS-CoV-2 infected subpopulation and the corresponding control subgroup. We also considered death after the 30 days since the infection as a competing risk. The overall results have been summarized in Fig. 3, which are further described below.

**Figure 3.**
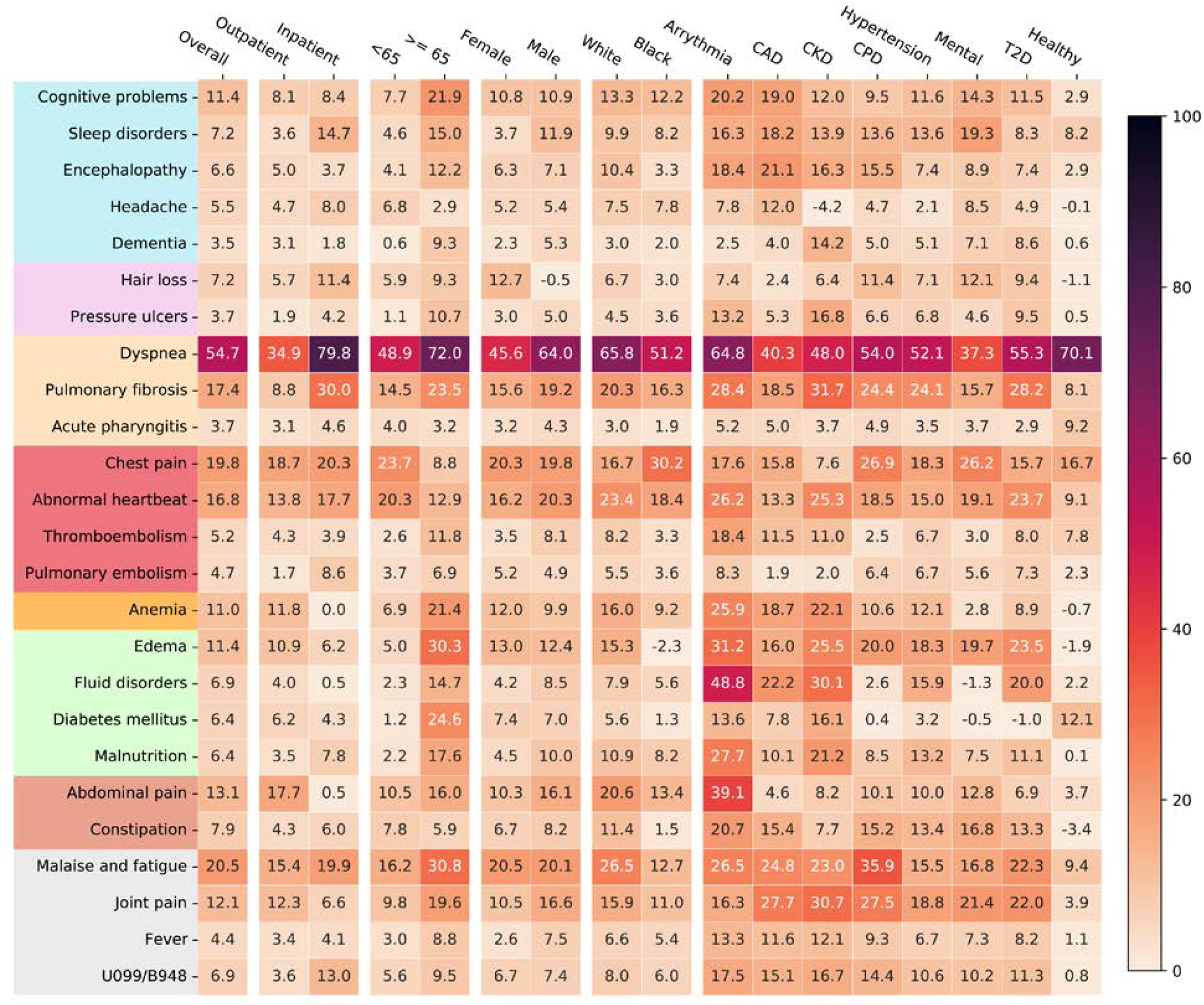
Adjusted excess cumulative incidence of post-acute sequelae of SARS-CoV-2 infection (PASC) in the INSIGHT cohort, from March 2020 to November 2021, stratified by the acute severity status, age groups, gender, race groups, and baseline pre-existing conditions. P-value < 3.6 × 10^−4^were used for selecting significant diagnoses. Different color panels represent different organ system, including (from top to bottom): nervous system, skin, respiratory system, circulatory system, blood forming organs, endocrine and metabolic, digestive system, genitourinary system, and general signs. CAD, coronary artery disease; CKD, chronic kidney disease; CPD, chronic pulmonary disease; T2D, diabetes type 2; Healthy: no documented pre-existing conditions and no PASC-like symptoms at baseline. Two ICD-10 diagnosis codes B948 (sequelae of other specified infectious and parasitic diseases) and U099 (post COVID-19 condition, unspecified) were also used to compare general post-acute sequelae of SARS-CoV-2 infection in different groups.

### Acute phase severity

General respiratory symptoms and signs (Fig. 3) demonstrated clear increasing burdens by settings (e.g., dyspnea from 34.9 excess cases per 1,000 patients compared to control patients in the outpatient setting to 79.8 in the inpatient setting). Other potential PASC diagnoses that followed the same trend included pulmonary fibrosis (8.8 to 30.0), sleep disorders (3.6 to 14.7), hair loss (5.7 to 11.4), and pulmonary embolism (1.7 to 8.6). We further investigated two PASC-related ICD-10 diagnosis codes, U099 (post COVID-19 condition, unspecified) and B948 (sequelae of other specified infectious and parasitic diseases), which also showed an increasing burden from 3.6 in the outpatient setting to 13.0 in the inpatient setting.

### Age groups

We partitioned patients into two groups according to their age (< 65 and ≥ 65). Potential PASC conditions that had the highest excess burden in <65 group were dyspnea, chest pain, abnormal heartbeat, malaise, and fatigue. Potential PASC conditions with highest excess burden in the ≥ 65 group included dyspnea, malaise and fatigue, edema, diabetes, anemia, cognitive problems, joint pain, malnutrition, and abdominal pain, among others; all of these conditions had a higher excess burden among those ≥ 65 compared to <65.

### Gender and race

Higher excess burdens in male patients included dyspnea, sleep disorders, malnutrition, and joint pain. Problems including hair loss and anemia demonstrated higher excess burdens for female patients. Black patients had higher excess burdens of chest pain than white patients.

### Baseline pre-existing conditions

Overall, we observed higher excess cumulative incidences in patients with any baseline pre-existing conditions (See Method) than patients without any assessed comorbidities or PASC-like symptoms (denoted as healthy patients). There were also varying excess burdens of different potential PASC conditions associated with patients with different pre-existing conditions. For example, patients with coronary artery disease (CAD) had higher burdens of cognitive problems, sleep disorders, and encephalopathy. Patients with chronic kidney disease (CKD) have higher burdens of pressure ulcers, diabetes, fluid disorders, and joint pains. Patients with chronic pulmonary disease (CPD) had higher burdens of malaise and fatigue, encephalopathy, hair loss, and chest pain than those without this condition at baseline.

### Comparison with the OneFlorida+ Cohort

To better understand the heterogeneity and commonality of potential PASC conditions over different populations, we replicated our analysis on the OneFlorida+ cohort and compared the adjusted hazard ratio of identified potential PASC diagnoses in the INSIGHT cohort versus the OneFlorida+ cohort. As shown in Fig. 4, overall higher adjusted hazard ratios were observed in the INSIGHT cohort compared to the OneFlorida+ cohort, indicating a generally higher risk of potential PASC conditions in INSIGHT than OneFlorida+. For certain PASC conditions the associated aHR values in the INSIGHT cohort exceed that in the OneFlorida+ cohort by more than 30%, such as malaise and fatigue, encephalopathy, sleep disorders, respiratory failure, pulmonary fibrosis, thromboembolism, anemia, and malnutrition. By calculating the excess burden of a set of potential PASC conditions identified from OneFlorida+ using the same criteria as INSIGHT (in Methods), we identified fewer “qualified” potential PASC conditions (Extended Data Fig. 2 and 3).

**Figure 4.**
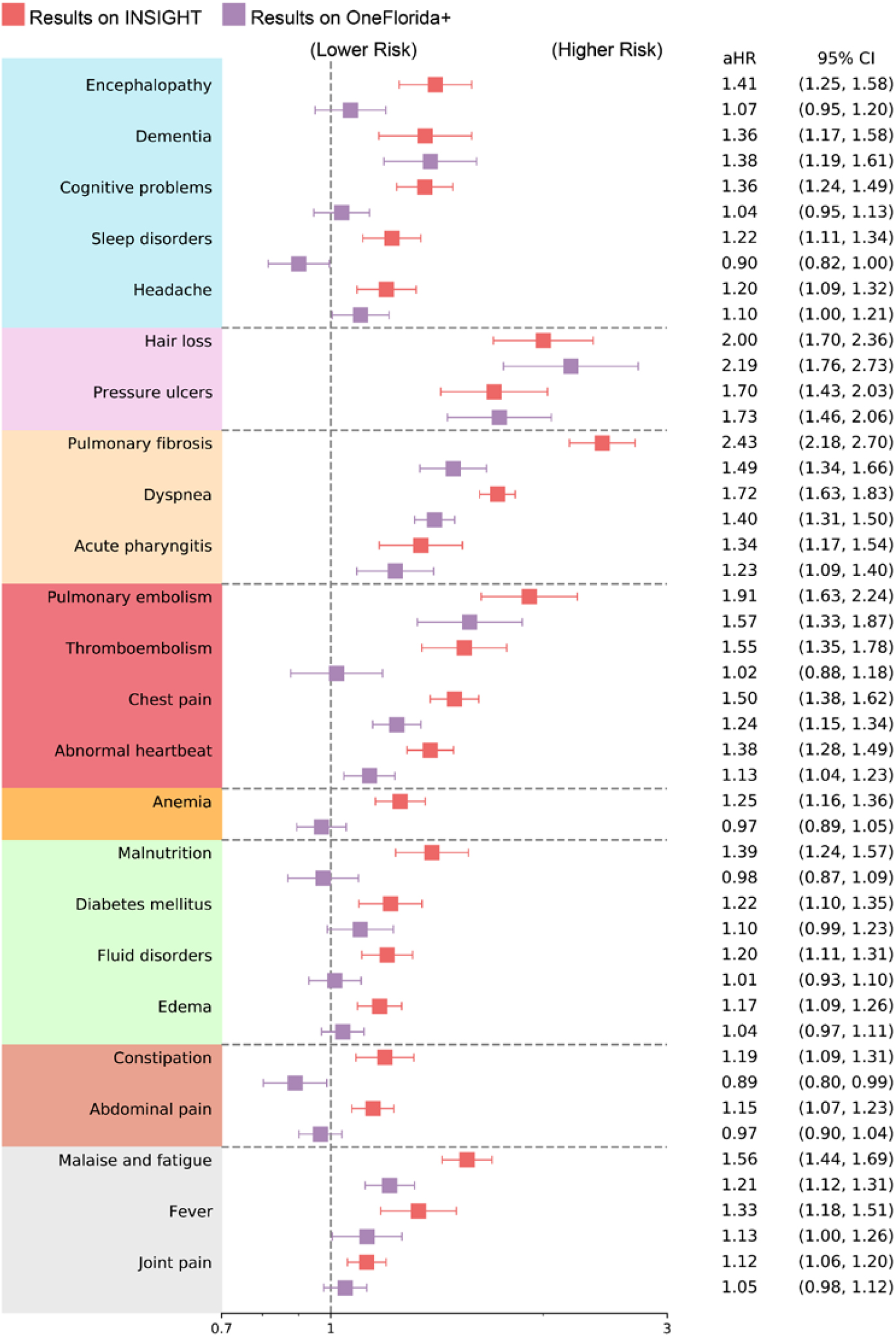
Comparison of the post-acute sequelae of SARS-CoV-2 risks in the INSIGHT cohort versus in the OneFlorida+ cohort, from March 2020 to November 2021. Adjusted hazard ratios were reported. The color panels represent different organ system, including (from top to bottom): nervous system, skin, respiratory system, circulatory system, blood forming organs, endocrine and metabolic, digestive system, and other general signs.

Some PASC conditions had similar risks in both the INSIGHT and OneFlorida+ cohorts, such as dementia, hair loss, pressure ulcers, acute pharyngitis, and diabetes mellitus (in Fig. 4). The stratified analysis results (Extended Data Fig. 2) also aligned with our observations in Fig.2, including significantly higher burdens of any PASC diagnoses in hospitalized patients in the acute phase, and malaise and fatigue, hair loss, dyspnea, and chest pain for female patients.

### Negative Controls

We employed negative outcome controls^19,20^ in both the INSIGHT and OneFlorida+ cohorts to rule out potential residual confounding. We examined the adjusted risk of a range of clinical outcomes (e.g., injury due to external causes and neoplasms-related outcomes) where no association was expected with COVID-19 based on prior knowledge. We emulated trials of negative outcomes following the same procedure as in screening potential PASC conditions and estimated the adjusted risk in both exposure groups. We found no significant association between any of the negative outcomes and SARS-CoV-2 infection after the acute phase as shown in Extended Data Table 1.

## Discussion

In this study, we developed a data-driven approach to identify a broad spectrum of clinical abnormalities (incident diagnoses and medication use) experienced by SARS-CoV-2 infected patients who survived beyond the first 30 days of the infection. The clinical EHRs from two large PCORnet clinical research networks, INSIGHT and OneFlorida+, were leveraged in our study to investigate the heterogeneity of potential PASC conditions over different patient populations. This differentiates our study from prior studies that focused on a specific patient population (e.g., Al-Aly et al. ^2^ focused on the US veteran population with 87.91% males). There are also several studies focusing on a more discrete set of potential PASC conditions such as mental health problems ^11,21^, cardiovascular problems ^10^, diabetes ^22^, kidney problems^23^ among others. Additionally, according to a recent systematic review^1^, most of these studies are small (less than 1,000 patients).

With a high-throughput causal inference pipeline, we identified a broad spectrum of diagnoses and medication use that exhibited higher adjusted hazard ratios and excess burdens in SARS-CoV-2 infected patients in the post-acute period compared to non-infected patients. These diagnoses and medications spanned a wide range of organ systems (Fig. 2), suggesting that PASC is a multi-organ disease. Diagnoses with high adjusted hazard ratios (aHRs) included respiratory problems (e.g., dyspnea and pulmonary fibrosis), dermatologic problems (e.g., hair loss and pressure ulcers), cardiovascular problems (e.g., pulmonary embolism, thromboembolism, chest pain, and abnormal heartbeat), nervous system problems (e.g., encephalopathy, dementia, and cognitive problems), and general symptoms (e.g., malaise, fatigue, fever, and joint pain). In addition to diagnoses, we also observed increased incident prescription risk in a diverse set of medications, including asthma drugs (e.g., vilanterol trifenatate and fluticasone furoate), cough drugs (e.g., dextromethorphan and benzonatate), anticoagulants (e.g., apixaban, heparin, and aspirin), diabetic drugs (e.g., insulin and metformin), drugs for constipation (e.g., magnesium hydroxide), drugs for vomiting (e.g., trimethobenzamide), pain medications (e.g., menthol, ibuprofen, and acetaminophen), drugs for treating skin problems (e.g., witch hazel and collagenase), and insomnia drugs (e.g., melatonin). These conditions and medications showed a higher incidence of diagnosis or use after infection than the non-infected control group, suggesting that these could be likely post-acute sequelae of SARS-CoV-2 infection (PASC) conditions.

We have also performed detailed stratified analyses on the adjusted excess burden of different potential PASC diagnoses over different groups defined by age, sex, race, acute severity of SARS-CoV-2 infection, and baseline comorbidity conditions. Our results showed that, in both the INSIGHT and OneFlorida+ cohorts, hospitalized patients demonstrated more excess cases of potential PASC diagnoses and medications (compared to non-infected controls) than non-hospitalized patients, especially for respiratory conditions. Older patients also had higher excess cases of PASC conditions than younger patients, as did female and non-white patients. These observations were consistent with prior studies.^2,18^ Patients with co-morbidities had a higher incidence and number of putative post-acute SARS-CoV-2 conditions. Furthermore, the distribution of post-acute conditions varied across distinct co-morbidities. We observed that dyspnea consistently showed the highest excess burden across all patients regardless of co-morbidity status. Patients with baseline cardiac problems (arrhythmia and coronary heart disease), type II diabetes, and chronic kidney disease (CKD) demonstrated higher burdens of a more diverse set of potential PASC conditions than other comorbidity groups. Patients without pre-existing conditions at baseline had higher dyspnea, chest pain, diabetes, malaise, and fatigue burdens compared with control patients.

We observed clear heterogeneity after replicating the same analysis to the OneFlorida+ cohort. Overall, aHRs on all potential PASC diagnoses identified from the INSIGHT cohort were higher than in the OneFlorida+ cohort. In particular, rates of pulmonary fibrosis and thromboembolism were 50% higher in the INSIGHT cohort than in the OneFlorida+ cohort. In addition, 25 diagnoses and 51 medications were identified as potential PASCs in the INSIGHT cohort compared to the 9 diagnoses and 9 medications identified in the OneFlorida+ cohort. Potential reasons accounting for this heterogeneity of PASC conditions include distinct patient characteristics and different periods of infections which could have led to differential use of therapeutics and vaccination that could alter the trajectory of PASC. The SARS-CoV-2 infected patients in the OneFlorida+ cohort were younger (median age 50 (34-64)) than those in the INSIGHT cohort (median age 55 (38-68)). Younger adults have been found to be at lower risk for PASC than older adults. Patients in the OneFlorida+ cohort were also much more socially disadvantaged on average. Their median ADI ranking value was almost four times higher than the median ADI of patients in the INSIGHT cohort. Disadvantaged social conditions can be associated with delayed or no care access and initiation of treatment for PASC conditions; therefore, OneFlorida+ patients might be less likely to present for care during a relatively short post-acute phase, leading to an undercount of potential PASC conditions and medication use in that population.

The treatment standard for COVID-19 evolved over time.^24,25^ For example, there was demonstrable higher use of corticosteroids in the Florida cohort compared with the NYC cohort. Patients who received timely and appropriate treatment for COVID-19 in the acute phase could be less likely to develop PASC in the post-acute phase. Early evidence showed that vaccinations for COVID-19 significantly reduced the likelihood of getting PASC conditions.^26^ It should be noted that NYC had a higher incident burden of SARS-CoV-2 infection prior to the widespread availability of vaccinations in December of 2020. In comparison, the OneFlorida+ had a high burden of incident SARS-CoV-2 infections after December 2020.

Our study has several strengths. First, this study examined PASC in a large population of general adult patients using a data-driven approach to identify a broad list of potential PASC. Existing studies with comparable sample sizes are Al-Aly et al. ^2^, which focused on a population of mostly male veterans, and Cohen et al. ^3^, which focused on older patients enrolled in a Medicare Advantage plan. Second, this study incorporated EHR data from two large-scale clinical research networks covering patients from distinct geographic regions in the US with very different characteristics, allowing us to highlight the heterogeneity of PASC manifestations in terms of diagnoses and medications over two different populations thereby improving generalizability. Third, from March 2020 to November 2021 (the enrollment period of our study), the US went through COVID-19 waves associated with different SARS-CoV-2 virus variants demonstrating different epidemiological and clinical characteristics. Our INSIGHT and OneFlorida+ cohorts contained robust patient populations in New York and Florida, representing the different waves of SARS-CoV-2 infected cases in the US. This temporal difference is another important factor accounting for the different observations from the two cohorts, in addition to their different demographic and geographic characteristics.

There are also several limitations. First, our study was based on observational data analysis, assignment to a particular exposure group was not randomized. However, we balanced high-dimensional hypothetical confounders and got consistent results from several negative outcome control analyses across two datasets, suggesting little confounding. Second, our study included the patient population from the NYC and Florida areas, which may not be representative of other geographical regions of the US or other countries. Third, the PASC is currently defined in the RECOVER protocols as “ongoing, relapsing, or new symptoms, or other health effects occurring after the acute phase of SARS-CoV-2 infection”. ^27^ Our study only studied incident events, and the worsening and relapsing conditions were left for future investigations. Fourth, the way these CCSR categories were defined may not reflect the actual co-occurring risk of the individual conditions contained in each in the context of PASC. In addition, our study period was from March 2020 to November 2021, which did not include patients infected during the phase dominated by the Omicron variants of SARS-CoV-2. Lastly, our analyses did not include information on vaccination status.

In conclusion, this study demonstrated that adult patients surviving beyond 30 days of their SARS-CoV-2 infection exhibited high incident risks and burdens across a broad range of conditions and signs. Our findings verified that PASC is a complex condition involving multiple organ systems. There was surprising geographic heterogeneity of PASC as well as patient sub-group heterogeneity. This study provides additional insights into our understanding of PASC and highlights the need for further research to support the diagnosis, prevention, and treatment of the post-acute sequelae of SARS-CoV-2 infection.

## Methods

### Data

This study used two large-scale de-identified real-world EHR datasets from the INSIGHT Clinical Research Network (CRN)^15^ and the OneFlorida+ CRN^16^. The INSIGHT CRN contained longitudinal clinical data of approximately 12 million patients in the New York City metropolitan area, and the OneFlorida+ CRN contained the EHR data of nearly 15 million patients from Florida and selected cities in Georgia and Alabama. The use of the INSIGHT data was approved by the Institutional Review Board (IRB) of Weill Cornell Medicine following NIH protocol 21-10-95-380 with protocol title: Adult PCORnet-PASC Response to the Proposed Revised Milestones for the PASC EHR/ORWD Teams (RECOVER). The use of the OneFlorida+ data for this study was approved under the University of Florida IRB number IRB202001831.

### High-throughput causal inference pipeline to identify post-acute sequelae of SARS-CoV-2 (PASC)

To systematically identify likely PASCs, we examined in total 596 incident diagnoses and medication use (Supplemental Table 1-2) in the SARS-CoV-2 infected patients from 31 days to 180 days after their acute infection. For each incident diagnostic category or medication use, we constructed an outcome-specific cohort including both SARS-CoV-2 infected patients and non-infected patients who did not have the corresponding diagnostic category or medication use at baseline and assessed its incident risk in the post-acute phase (see Fig. 1 for a graphical illustration of our pipeline). Similar to trial emulation using real-world data^27^, we evaluated the impact of SARS-CoV-2 infection (as exposures) in the post-acute period using each target outcome, leading to 596 independent trials. Of note, we scaled up standard trial emulation with only one specific outcome to our data-driven high-throughput hypotheses generation setting described as follows. We further summarized the key protocol components of these target trials and their high-throughput emulations in Extended Data Table 2.

### Eligibility criteria and exposure strategies

We included patients with at least one SARS-CoV-2 polymerase-chain-reaction (PCR) or antigen laboratory test between March 01, 2020, and November 30, 2021, for both cohorts. Other eligibility criteria included an age of at least 20 years old, at least one diagnosis code within three years to seven days prior to the index date (referred to as the baseline period), and at least one diagnosis code from 31 days to 180 days after the index date (referred to as the post-acute phase or follow-up period), to ensure that patients were connected to the healthcare system and were being observed during the study period. Two exposure groups were the SARS-CoV-2 infected group and the non-infected group. The SARS-CoV-2 infected group included patients with a positive SARS-CoV-2 PCR or antigen laboratory test. The index date of the infected group was defined as the date of the first documented positive PCR or antigen test. The non-infected group included patients whose SARS-CoV-2 PCR or Antigen tests were all negative throughout the entire study period with no documented COVID-19 related diagnoses at any time. The index date for patients in the non-infected group was defined as the date of the first negative PCR or antigen test. See the Supplementary Table 1 for the list of LOINC laboratory codes and ICD-10 diagnosis codes used for building patient cohorts.

### Group assignment and baseline covariates

We assigned patients to the two exposure groups (i.e., the SARS-Cov-2 infected group and the non-infected group) according to their baseline eligibility criteria and the exposure strategies. Patients in the two exposure groups were assumed exchangeable after adjusting for high-dimensional baseline covariates as hypothetical confounders. The collected baseline covariates included age (categorized into 20-39 years, 40-54 years, 55-4 years, 65-74 years, 75-84 years, 85 years and older), gender (female, male, other/missing), race (Asian, Black or African American, White, other, missing), ethnicity (Hispanic, not Hispanic, other/missing). The national-level area deprivation index (ADI) was used to capture the socioeconomic disadvantage of patients’ residential neighborhood^17^. We used 9-digit zip code to link to the national ADI percentiles (ranked from 1 to 100). We imputed missing ADI value with median ADI per site. Healthcare utilization was measured as the number of inpatient, outpatient, and emergency encounters (0 visit, 1 or 2 visits, 3 or o visits, 5+ visits for each encounter type) respectively. In addition, periods (March 2020 – June 2020, July 2020 – October 2020, November 2020 - February 2021, March 2021 – June 2021, July 2021 – November 2021) of the index date were used to account for potentially different stages of the pandemic. We also collected a wide range baseline comorbidities based on a tailored list of the Elixhauser comorbidities^28^ and related drug categories, including alcohol abuse, anemia, arrythmia, asthma, cancer, chronic kidney disease, chronic pulmonary disorders, cirrhosis, coagulopathy, congestive heart failure, chronic obstructive pulmonary disease, coronary artery disease, dementia, diabetes type 1, diabetes type 2, end stage renal disease on dialysis, hemiplegia, HIV, hypertension, hypertension and type 1 or 2 diabetes diagnosis, inflammatory bowel disorder, lupus or systemic lupus erythematosus, mental health disorders, multiple sclerosis, Parkinson’s disease, peripheral vascular disorders, pregnant, pulmonary circulation disorder, rheumatoid arthritis, seizure/epilepsy, severe obesity (BMI>=40 kg/m^2^), weight loss, Down’s syndrome, other substance abuse, cystic fibrosis, autism, sickle cell, corticosteroid drug prescriptions, immunosuppressant drug prescriptions. Patients were defined as having a condition if they had at least two corresponding diagnoses documented during the baseline period.

### Follow-up period

We followed each patient from 31 days after his/her index date until the day of the first target outcome, documented death, loss of follow-up in the database, 180 days after the baseline, or the end of our observational window (December 31, 2021), whichever came first.

### Diagnosis categories for screening potential PASC conditions

We examined an initial list of potential adult PASC diagnostic outcomes for screening, which contained 137 diagnostic categories. A team of clinicians built our initial screening list based on the Clinical Classifications Software Refined (CCSR) v2022.1 covering all the 66,534 ICD-10-CM Diagnoses, removed codes that cannot be attributed to COVID-19 (e.g., HIV, tuberculosis, infection by non-COVID causes, neoplasms, injury due to external causes), and systematically added parent codes (e.g., the first 3-digits of ICD-10 codes) of potential PASC diagnosis codes. The full list of our investigative diagnosis codes is provided in Supplementary Table 2 and includes 6466 codes.

### Medications for screening potential PASC conditions

We examined an initial list of potential adult PASC medication outcomes for screening, which contained 459 drug categories classified by their active ingredients. We collected real-world drug prescription data from our EHR datasets, mapped drugs into their active ingredients, and selected drug ingredients prescribed for at least 100 patients in the COVID-19 positive group, which led to 434 active drug ingredients. We further considered another 25 categories of medications used during the course of treatment for COVID-19, including anti-platelet therapy, aspirin, colchicine, corticosteroids, dexamethasone, and heparin, which were potentially identified by both prescription records and procedure records.

### Causal contrasts of PASC outcomes’

Adjusted hazard ratio and excess burden for each incident PASC diagnosis or medication were calculated in the follow-up period.

### High-throughput screening pipeline for PASC

We systematically examined the 137 diagnosis categories and 459 medication ingredients using our pipeline shown in Fig. 1.

### Statistical analyses for high-throughput hypotheses generation

#### Inverse propensity score weighting confounding control

We built a propensity score (PS) model—the probability of assignment of a particular exposure group conditioned on baseline covariates—for each target outcome. Based on the estimated PS values, we then used stabilized inverse propensity score weighting (IPTW) ^29^ to re-weight patients in exposure and control groups, aiming to balance the two groups on baseline covariates after re-weighting. If we use *X, Z* to represent the observed baseline covariates and the assignment of exposure (*Z* = 1) and control groups (*Z* = 0), the PS is defined as *P*_*θ*_ (*Z* = 1|*X)* and the stabilized IPTW is 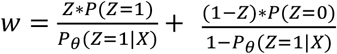. We used standardized mean difference (SMD) to quantify the goodness-of-balance of covariates over two groups 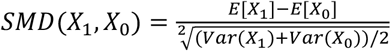and used *SMD* < 0.*1* as the threshold for balancing diagnostics. The SMD was calculated before and after IPTW re-weighting, and the results are provided in Supplementary Table 3 and 4.

We used regularized logistic regression for PS calculation, with the optimal regularization parameters determined through grid search. We have shown in our previous study that a better PS model can be selected by considering both the goodness-of-balance performance and the goodness-of-fit performance. Here, we designed a cross-validation pipeline for PS model training, selection, and validation as detailed in Extended Data Table 3 based on our prior work^30^, which can achieve better goodness-of-balance performance compared with the PS model selected by other machine learning model selection strategies in trial emulations.

#### Statistical analysis for causal contrasts

The adjusted hazard ratio (aHR) was estimated by Cox proportional hazard model with the abovementioned stabilized IPTW weights. The cumulative incidence was estimated by the Aalen-Johansen model^31^ considering death to be a competing risk for the target outcomes.

#### Subgroup analysis and negative outcome controls

The subgroup analysis was conducted by stratifying patients (in both SARS-CoV-2 infected and non-infected groups) by their age, race, gender, the severity of acute infection (outpatient or inpatient) and pre-existing conditions. We also included a population with no documented pre-existing conditions or PASC-like symptoms at baseline, denoted as healthy. To explore the possible existence of residual confounding, we estimated the adjusted hazard ratio of non-PASC outcomes following negative outcome control framework^19,20^.

#### Screening criteria for likely PASC conditions

To reduce the chance of false positive discovery, we adhered to the following screening criteria: Only diagnoses and medications with 1) adjusted hazard ratios larger than 1, 2) P-value smaller than 3.6 × 10^−4^(for diagnoses, significance level corrected by Bonferroni method for multiple testing) and 1.4 × 10^−4^(for medications) will be retained as potential PASCs. Further, we required a minimum number of PASC events that appeared (at least 100 times on INSIGHT and 63 times on OneFlorida+) in the post-acute period of COVID-19 patients. We also considered Holms’ method for correcting the P-value under the high-throughput multiple-test setting, and both correction methods gave a consistent conclusion.

## Code availability

For reproducibility, our codes are available at https://github.com/calvin-zcx/pasc_phenotype. We used Python 3.9, python package lifelines-0.2666 for survival analysis, and scikit-learn-0.2318 for machine learning models.

## Data Availability

The INSIGHT data can be requested through https://insightcrn.org/. The OneFlorida+ data can be requested through https://onefloridaconsortium.org. Both the INSIGHT and the OneFlorida+ data are HIPAA-limited. Therefore, data use agreements must be established with the INSIGHT and OneFlorida+ networks.

## Acknowledgment

This research was funded by the National Institutes of Health (NIH) Agreement OTA HL161847-01 (contract number EHR-01-21) as part of the Researching COVID to Enhance Recovery (RECOVER) research program.

## Extended Data Figures and Tables

**Extended Data Fig. 1.**
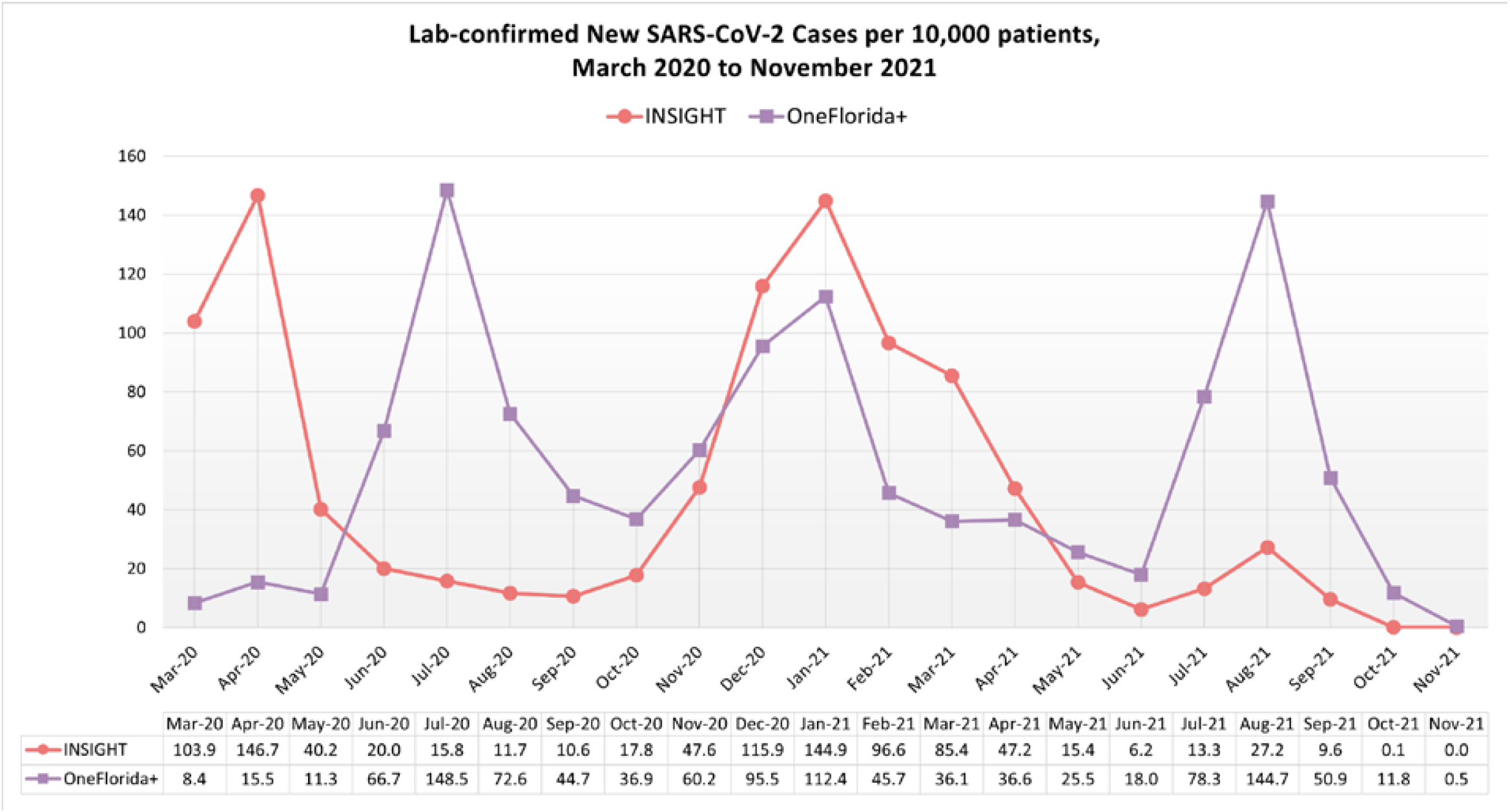
Lab-confirmed New SARS-CoV-2 Cases Per 10,000 patients in the INSIGHT and OneFlorida+ cohorts, from March 2020 to November 2021.

**Extended Data Fig 2.**
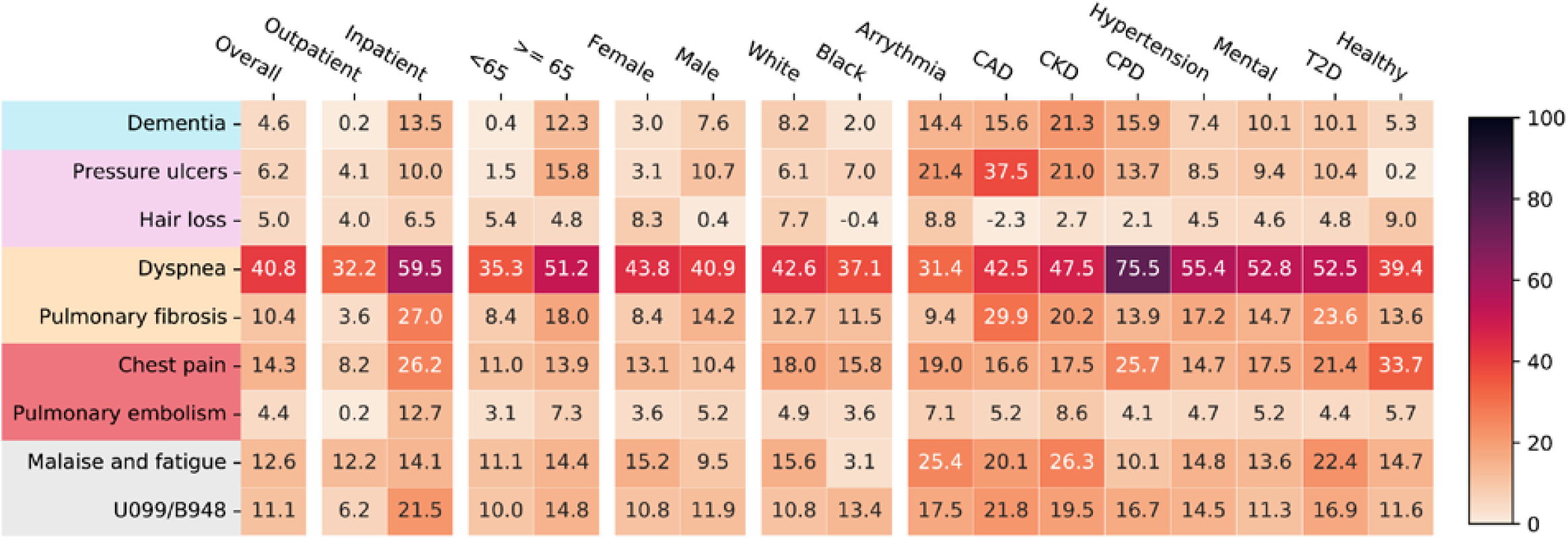
Adjusted excess cumulative incidence of post-acute sequelae of SARS-CoV-2 infection (PASC) in the OneFlorida+ cohort, March 2020 – November 2021, stratified by the acute severity status, age groups, gender, race groups, and pre-existing conditions. P-value < 3.6 × 10^−4^were used for selecting significant diagnoses. Different color panels represent different organ system, including (from top to bottom): nervous system, skin, respiratory system, circulatory system, musculoskeletal system, and general signs. CAD, coronary artery disease; CKD, chronic kidney disease; CPD, chronic pulmonary disease; T2D, diabetes type 2.

**Extended Data Fig 3.**
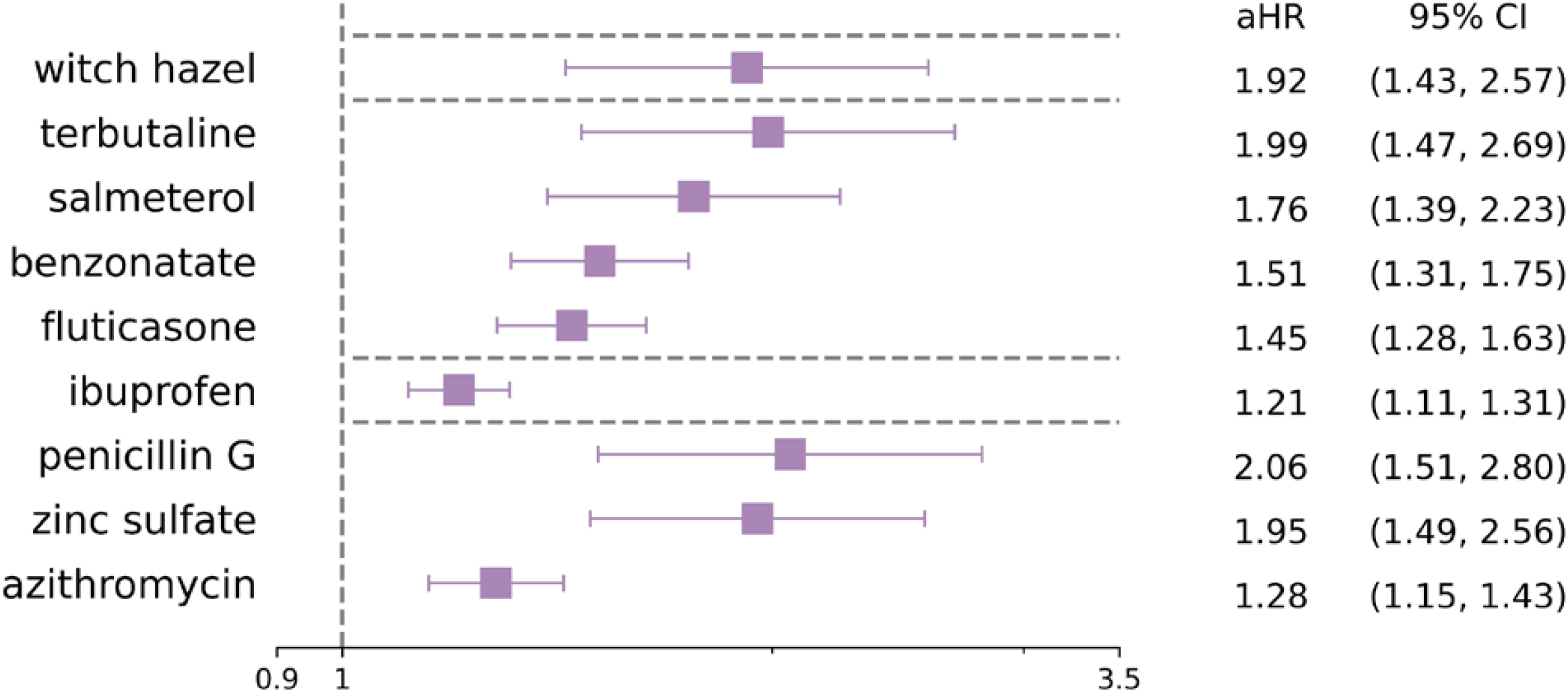
Adjusted hazard ratio of identified incident medications for PASC in the OneFlorida+ cohort. Bonferroni corrected significance level 1.4 × 10^−4^was used for selecting significant medications.

**Extended Data Table 1.** Results of negative outcome control in both the INSIGHT and OneFlorida+ cohorts, March 2020– November 2021.

|  | Adjusted Hazard Ratio (95% Confidence Interval)<sup>a</sup> | P-value<sup>b</sup> | No. of Covid-19 Positive patients | No. of patients in control group<sup>c</sup> | Number of events in case group | Number of events in control group | No. of unbalanced covariates | No. of unbalanced covariates after re-weighting<sup>a</sup> |
| --- | --- | --- | --- | --- | --- | --- | --- | --- |
| **Negative Outcomes** |  |  |  |  |  |  |  |  |
| **INSIGHT** |  |  |  |  |  |  |  |  |
| Accidental Injuries | 1.02 (0.84, 1.23) | 0.87 | 33,898 | 33,898 | 347 | 248 | 26 | 0 |
| Benign Neoplasms | 0.93 (0.84, 1.03) | 0.14 | 30,461 | 30,461 | 1030 | 1228 | 26 | 0 |
| **OneFlorida+** |  |  |  |  |  |  |  |  |
| Accidental Injuries | 0.99 (0.88, 1.12) | 0.92 | 18,967 | 18,967 | 661 | 599 | 10 | 0 |
| Benign neoplasms | 0.93 (0.82, 1.05) | 0.23 | 18,729 | 18,729 | 585 | 693 | 10 | 0 |

- Outcomes were ascertained from day 30 after the SARS-CoV-2 infection. The adjusted hazard ratios were computed at 180 days after the SARS-CoV-2 infection by adjusting high-dimensional baseline variables the same as in the screening sequelae as discussed in the Method section. We used inverse propensity treatment weights to do re-weight. All the baseline covariates were balanced in term of standardized mean difference < 0.1. - Large P-value (> 0.05) indicates no significant association between SARS-CoV-2 infection and outcomes were found - The number of patients in the control group were randomly selected from all Covid-19 negative patients with the same number as patients in the case group.

**Extended Data Table 2.** Specifications and High-throughput emulations of target trials (a large number of hypothetical randomized controlled clinical trials to answer a large number of causal questions of interests) to screening potential Post-Acute Sequelae of SARS-CoV-2 infected (PASC) using INSIGHT Electronic Health Records in New York City and OneFlorida+ EHRs in Florida (March 2020 – November 2021).

| Protocol component | Specifications of Target Trials | High-throughput target trial emulation |
| --- | --- | --- |
| Eligibility criteria | • Age $\geq 20$ years at index, and no upper age limit, between March 1, 2020, and November 30, 2021.<br>• Patients without any positive SARS-CoV-2 polymerase-chain-reaction (PCR) or Antigen test, or COVID-19 diagnoses before index<br>• Use of the INSIGHT health care system, defined as at least one encounter with any diagnoses, in the past 3 years to 7 days before index.<br>• Use of the INSIGHT health care system, defined as at least one encounter with any diagnoses, 31 days to 180 days after index in the follow-up period, indicates being alive after potential COVID-19 acute phase and at least 31 days of potential follow-up.<br>• Known documented SARS-CoV-2 PCR/Antigen lab positive or negative results<br>• (High-throughput scenario for PASC) No history of target post-acute sequelae of COVID-19 in the past 3 years to 7 days before index.<br>• (High-throughput scenario for PASC medications) No history of target medication usage in the past 1 year to 7 days before index. | Same as for the target trials. We identified the SARS-CoV-2 infected PCR/Antigen tests using the laboratory results table, and the COVID-19 diagnoses using ICD-10 codes in patients' diagnosis table in the INSIGHT EHR system or OneFlorida+ EHR system, following the PCORnet data model |
| Exposure strategies | • Exposure group: Infection of SARS-CoV-2 and the SARS-CoV-2 PCR/Antigen tested positive<br>• Control group: No infection of SARS-CoV-2, and the SARS-CoV-2 PCR/Antigen tests keeping negative in the follow-up period | • Same as for the target trial.<br>• SARS-CoV-2 infected cases were defined as patients who had any SARS-CoV-2 positive PCR/Antigen test, and the baseline date, or the index date, is defined as the date of the first documented positive SARS-CoV-2 PCR/Antigen test.<br>• Non-infected controls were defined as patients whose SARS-CoV-2 PCR/Antigen tests were all negative, and there were no COVID-19 related diagnoses documented anytime. The baseline date, or the index date, is defined as the date of the first documented SARS-CoV-2 PCR/Antigen test. |
| Group assignment | Individuals are randomly assigned to an exposure strategy at baseline and are aware of the assigned exposure strategy. | We classified patients into different exposure groups according to their baseline eligibility criteria and the exposure strategies. We assumed that the exposure group and the control group were exchangeable by adjusting for high-dimensional baseline, including age, gender, race, ethnicity, social-economic status, hospital utilization history, time period of infection, baseline comorbidities, history prescriptions, etc. |
| High-throughput Outcomes | • 137 potential post-acute sequelae of COVID-19 (PASC)<br>• 459 categories of drugs due to post-acute sequelae of COVID-19<br>Hospitalization due to PASC<br>ICU admission due to PASC<br>Death due to PASC (defined as death after 31 days of SARS-CoV-2 infection) | Same as for the target trials. |
| Follow-up | We followed each patient from his/her baseline day until the day of the outcome of interest, death, 180 days after baseline, or the end of the study period (November 30, 2021), whichever happens first. | Same as for the target trial. |
| Causal contrasts | Excess risk of newly-onset post-acute sequelae of COVID-19 against baseline incidence. | Same as for the target trial. |
| High-throughput trials for screening PASC | For each target PASC outcome, we conducted a corresponding target trial among which patients were free of the target PASC outcome at baseline and no history of target outcome before baseline. The number of potential PASCs are large and thus the number of target trials are large. | We emulated 137 trials for potential PASC diagnosis outcomes, and 459 trials for potential PASC medication outcomes. For each emulated trial, the exposure group consisted of eligible SARS-CoV-2 infected patients without a history of target outcome at baseline. The control group were built from COVID-19 negative patients who were also without any history of target outcome at baseline. The ratio of the number of patients in the exposure group and the control group was 1:5. |
| Statistical analysis | Cumulative incidence (risk) curves and estimates of 18-months risk, and hazard ratio (HR) between two exposure groups.<br>Subgroup analyses by baseline age, race, gender, and severity of acute phase of COVID-19. | Same as for the target trial.<br>• We used inverse propensity of treatment re-weighting (IPTW) to adjust for high-dimensional baseline covariates. We used L2-norm logistic regression for propensity score calculation, and the best PS model was selected in 10-fold cross-validations with respect to both goodness-of-balance and goodness-of-fit.<br>• Adjusted Cox proportional hazard model, Kaplan-Meier estimator, and Aalen-Johansen estimator were used.<br>• The p-value was corrected by Bonferroni method for multiple tests of diagnoses and medications separately.<br>• Potential PASCs were selected according to the number of patients with target outcome in the real-world data (>200), the corrected P-value ($<0.05/\text{number of trials}$), and the hazard ratio ($>1$) |

Abbreviations: PCORnet, the National Patient-Centered Clinical Research Network; PASC, Post-Acute Sequelae of COVID-19; PCR, polymerase chain reaction; HR, hazard ratio; IPTW, inverse propensity of treatment re-weighting

**Extended Data Table 3.**
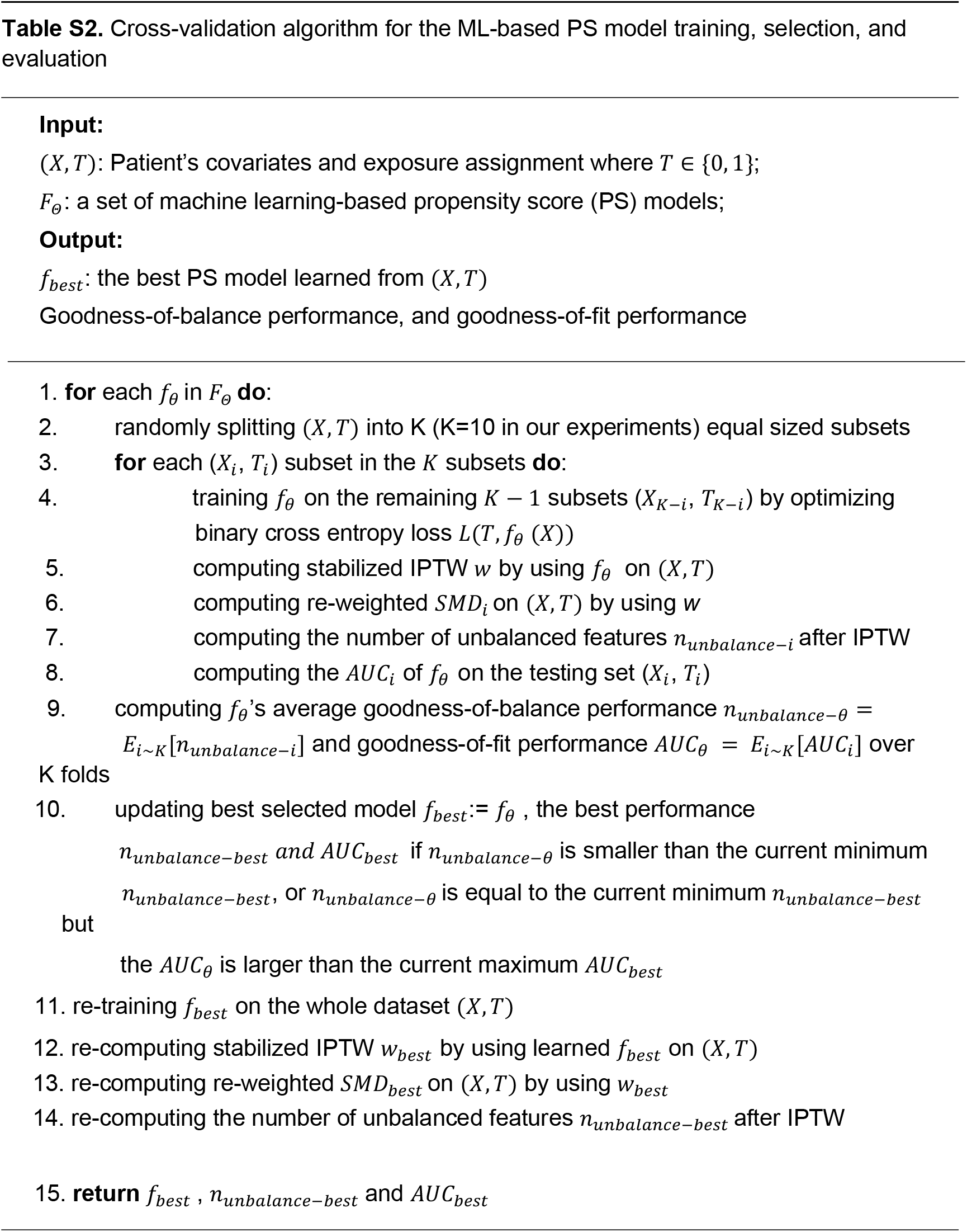
Cross-validation algorithm tailored for our machine learning-based propensity score calculation for each emulated trial.

## Supplementary Information

- **Supplementary Table 1**. COVID-19 Phenotyping Lab LOINC codes and Diagnosis ICD10 codes
- **Supplementary Table 2**. PASC Adult Diagnostic List for Screening
- **Supplementary Table 3**. Balance Diagnostics of Discovered PASC Specific Cohorts on INSIGHT, NYC
- **Supplementary Table 4**. Balance Diagnostics of Discovered PASC Specific Cohorts on OneFlorida, Florida

